## Supplementary figures and images for "Automatic time in bed detection from hip-worn accelerometers for large epidemiological studies: The Tromsø Study"

### S1 Fig.

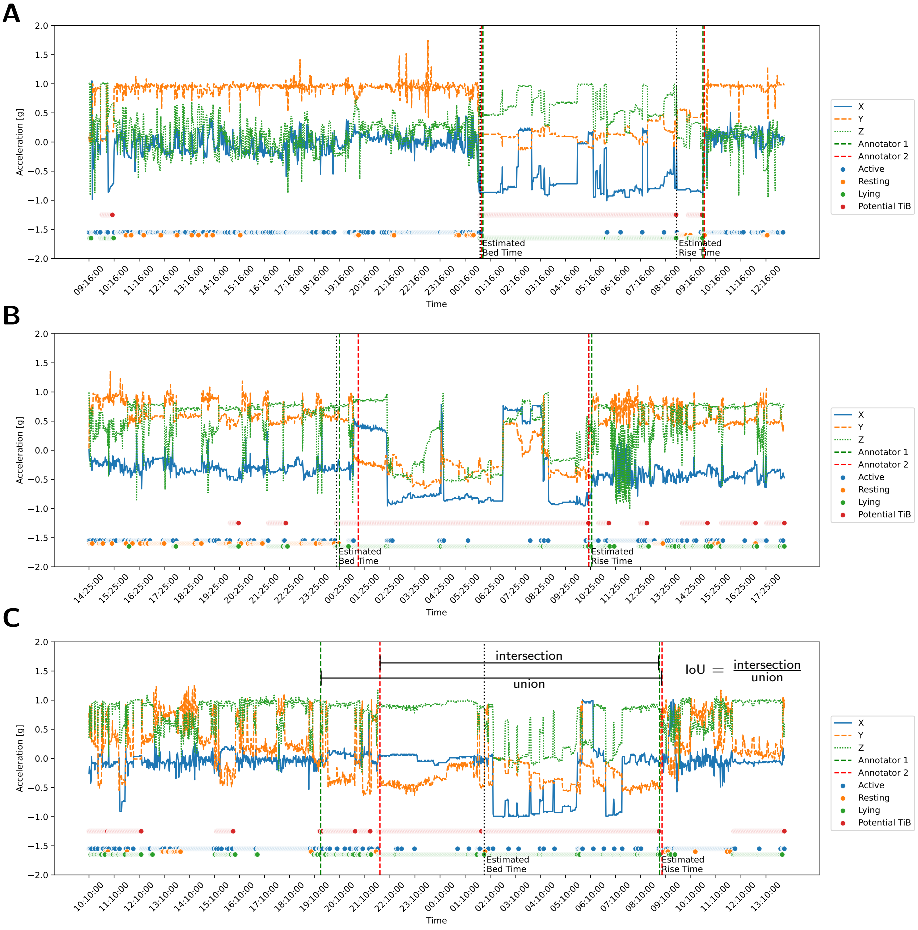

### S3 Fig.

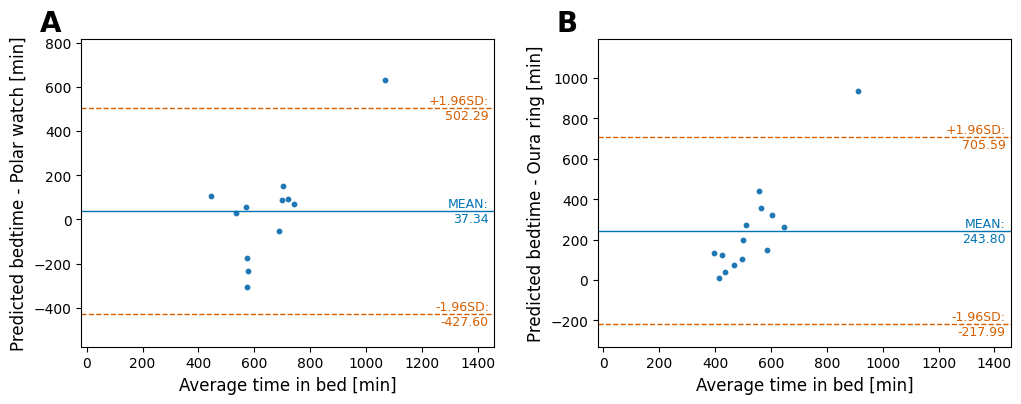

### S4 Fig.

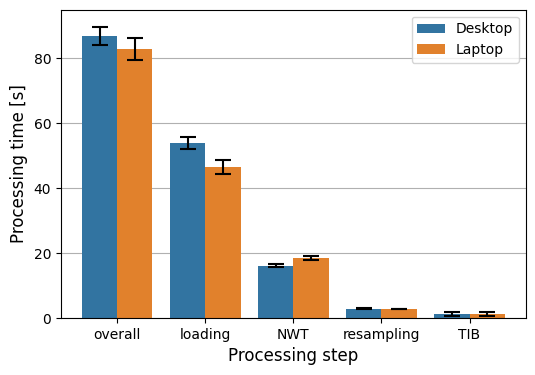
