## Supplementary material for "Automatic time in bed detection from hip-worn accelerometers for large epidemiological studies: The Tromsø Study": S2 Table.

| Model | Layers | LSTM cells 1 | 2 | 4 | 8 | 16 | 32 | 64 | 128 |
|---|---|---|---|---|---|---|---|---|---|
| Uni-LSTM | 1 | .90 ± .00 | .92 ± .01 | .91 ± .01 | .92 ± .01 | .92 ± .01 | .92 ± .01 | .92 ± .01 | .92 ± .01 |
| Uni-LSTM | 2 | .90 ± .01 | .89 ± .01 | .91 ± .01 | .92 ± .01 | .92 ± .01 | .92 ± .01 | .92 ± .00 | .92 ± .00 |
| Uni-LSTM | 4 | .79 ± .05 | .89 ± .01 | .92 ± .01 | .92 ± .01 | .92 ± .00 | .93 ± .00 | .94 ± .00 | .92 ± .01 |
| Bi-LSTM | 1 | .91 ± .01 | .91 ± .01 | .92 ± .01 | .94 ± .01 | .94 ± .01 | .94 ± .01 | .94 ± .01 | .94 ± .01 |
| Bi-LSTM | 2 | .91 ± .01 | .92 ± .01 | .94 ± .01 | .94 ± .01 | .94 ± .01 | .95 ± .01 | .95 ± .01 | .95 ± .01 |
| Bi-LSTM | 4 | .92 ± .01 | .93 ± .01 | .94 ± .01 | .95 ± .01 | .95 ± .01 | .95 ± .00 | .95 ± .00 | .95 ± .01 |

Uni-LSTM: Unidirectional LSTM; Bi-LSTM: Bidirectional LSTM
