## Supplementary material for "Automatic time in bed detection from hip-worn accelerometers for large epidemiological studies: The Tromsø Study": S5 Table.

| | Accuracy | F1 Score | Recall | Precision | Sensitivity | Specificity | Total TiB | Total predicted TiB |
|---|---|---|---|---|---|---|---|---|
| All days | 0.920 (0.130) | 0.904 (0.141) | 0.966 (0.097) | 0.875 (0.188) | 0.966 (0.097) | 0.894 (0.200) | 493.471 (93.179) | 576.822 (213.178) |
| w/o outlier predictions | 0.933 (0.099) | 0.917 (0.115) | 0.969 (0.075) | 0.890 (0.162) | 0.969 (0.075) | 0.914 (0.152) | 494.305 (91.133) | 560.375 (168.907) |
| w/o outlier predictions and NWT days | 0.935 (0.098) | 0.917 (0.114) | 0.969 (0.076) | 0.892 (0.161) | 0.969 (0.076) | 0.916 (0.150) | 491.607 (94.666) | 555.291 (169.046) |
