## Supplementary material for "Automatic time in bed detection from hip-worn accelerometers for large epidemiological studies: The Tromsø Study": S6 Table.

| | All days | | w/o outlier predictions | | w/o outlier predictions and NWT days | |
|---|---|---|---|---|---|---|
| | Labeled | Predicted | Labeled | Predicted | Labeled | Predicted |
| 0.01 | 287.60 | 255.44 | 321.04 | 279.20 | 275.71 | 273.04 |
| 0.10 | 392.00 | 392.00 | 392.20 | 392.60 | 390.00 | 388.00 |
| 0.25 | 435.00 | 457.00 | 435.00 | 457.00 | 431.00 | 449.00 |
| 0.50 | 490.00 | 530.00 | 489.00 | 529.00 | 488.00 | 523.00 |
| 0.75 | 543.00 | 623.00 | 543.00 | 619.00 | 541.00 | 613.00 |
| 0.90 | 604.00 | 807.00 | 601.00 | 764.20 | 605.36 | 758.60 |
| 0.99 | 721.72 | 1440.00 | 721.96 | 1150.12 | 726.88 | 1150.12 |
